# Utility of surveillance data for planning for dengue elimination in Yogyakarta, Indonesia: a scenario tree modelling approach

**DOI:** 10.1101/2023.03.14.23287232

**Authors:** Melanie Bannister-Tyrrell, Alison Hillman, Citra Indriani, Riris Andono Ahmad, Adi Utarini, Cameron P. Simmons, Katherine L. Anders, Evan Sargeant

## Abstract

**Introduction:** Field trials and mathematical modelling studies suggest that elimination of dengue transmission may be possible through widespread release of *Aedes aegypti* mosquitoes infected with the insect bacterium *Wolbachia pipientis* (*w*Mel strain), in conjunction with routine dengue control activities. This study aimed to develop a modelling framework to guide planning for the potential elimination of locally-acquired dengue in Yogyakarta, a city of almost 400,000 people in Java, Indonesia.

**Methods:** A scenario tree modelling approach was used to estimate the sensitivity of the dengue surveillance system (including routine hospital-based reporting and primary-care based enhanced surveillance), and time required to demonstrate elimination of locally-acquired dengue in Yogyakarta city, assuming the detected incidence of dengue decreases to zero in the future. Age and gender were included as risk factors for dengue, and detection nodes included the probability of seeking care, probability of sample collection and testing, diagnostic test sensitivity, and probability of case notification. Parameter distributions were derived from health system data or estimated by expert opinion. Alternative simulations were defined based on changes to key parameter values, separately and in combination.

**Results:** For the default simulation, median surveillance system sensitivity was 0.131 (95% PI 0.111 – 0.152) per month. Median confidence in dengue elimination reached 80% after a minimum of 13 months of zero detected dengue cases and 90% confidence after 25 months, across different scenarios. The alternative simulations investigated produced relatively small changes in median system sensitivity and time to elimination.

**Conclusion:** This study suggests that with a combination of hospital-based surveillance and enhanced clinic-based surveillance for dengue, an acceptable level of confidence (80% probability) in the elimination of locally acquired dengue can be reached within 2 years. Increasing the surveillance system sensitivity could shorten the time to first ascertainment of elimination of dengue and increase the level of confidence in elimination.

**Key messages:** *What is already known on this topic - summarise the state of scientific knowledge on this subject before you did your study and why this study needed to be done:* The incidence of dengue, a mosquito-borne viral disease, has increased worldwide in recent decades. However, a novel vector control intervention based on the release of *Aedes aegypti* vectors infected with the *w*Mel strain of *Wolbachia pipientis* bacteria, which inhibits arboviral infection and transmission in mosquitoes, has been shown to substantially decrease the incidence of dengue. A randomised controlled trial in Yogyakarta city, Indonesia, demonstrated a 77% reduction in dengue incidence in areas treated with the *Wolbachia* intervention. Field-based and modelling studies have predicted that the *Wolbachia* intervention could reduce dengue transmission to the point of elimination of locally-acquired dengue. The feasibility of elimination of dengue as a public health problem through city-wide deployment of *Wolbachi*a intervention, complemented by other vector control measures and enhanced clinical surveillance, is being tested in Yogyakarta city, Indonesia, as part of an additional study. For any planned infectious disease elimination program, demonstrating that elimination has been achieved requires effective surveillance for dengue and robust statistical methods. The aim of this study was to develop a scenario tree modelling framework to guide planning for the potential elimination of locally acquired dengue in Yogyakarta city, Indonesia.

*What this study adds - summarise what we now know as a result of this study that we did not know before:* This study constructed a scenario tree model to represent the dengue surveillance system in Yogyakarta city, including hospital-based and clinic-based surveillance. The model was parameterised with risk factors for dengue, as well as the steps in the surveillance pathway, from healthcare seeking to notification of a positive case. This study suggests that with a combination of hospital-based surveillance and enhanced clinic-based surveillance for clinical dengue cases, an acceptable level of confidence (80% probability) in the elimination of locally acquired dengue in Yogyakarta city can be reached within 2 years of the last detected case. This suggests that routine clinic-based and hospital-based passive surveillance for dengue is sufficiently sensitive to plan for and confirm elimination of local dengue transmission in Yogyakarta city.

*How this study might affect research, practice or policy - summarise the implications of this study:* The findings suggest that pragmatic demonstration of the elimination of locally-acquired dengue can be reached in a shorter timeframe than the alternative approach of convening an independent expert review panel, as commonly used for other infectious disease elimination programs. The scenario tree modelling framework offers a replicable, robust method for planning and assessing progress towards elimination of dengue as a public health problem following full implementation of the *Wolbachia* intervention and enhanced clinic-based dengue surveillance in Yogyakarta city, Indonesia. The framework can be readily expanded and adapted to different geographic settings where the *Wolbachia* intervention is also being implemented to control and eliminate dengue.

## Introduction

Dengue is an acute viral syndrome caused by infection with any of four dengue virus serotypes transmitted by the bite of infected *Aedes* mosquito vectors, of which *Ae. aegypti* is the primary vector [1]. The incidence of dengue has increased substantially in recent decades, with an eight-fold increase in cases reported to the World Health Organization between 2000 and 2019 [2]. Though most dengue infections are asymptomatic, an estimated 50 – 100 million symptomatic cases occur per year, which are associated with approximately 1.14 million disability-adjusted life years lost [3]. The World Health Organization designated dengue as one of the top 10 global health threats in 2019 [4].

Dengue prevention and control has historically been based on vector control measures, especially control and removal of vector breeding sites. These methods have had limited effectiveness in controlling dengue as a public health problem in most endemic settings [1]. An innovative dengue control intervention has been developed based on the release of *Ae. aegypti* vectors infected with the *w*Mel strain of *Wolbachia pipientis* bacteria, which inhibits arboviral infection and transmission (hereafter the *Wolbachia* intervention). The *Wolbachia* intervention for dengue control is described elsewhere [5–9]. A cluster randomised trial of the *Wolbachia* intervention in Yogyakarta city, Indonesia, reported a 77.1% reduction in the odds of dengue in the intervention clusters compared to control clusters [10]. This builds on earlier evidence of the effectiveness of the *Wolbachia* intervention in substantially decreasing dengue incidence. In northern Queensland, Australia, clinical dengue incidence was reduced by 96% following a series of *Wolbachia* mosquito releases, and *Wolbachia* was sustained at high levels in local *Ae aegypti* populations more than eight years after release [11]. Mathematical modelling studies have predicted that the *w*Mel-infected *Ae. aegypti* intervention could reduce the effective reproduction number for dengue to below one, leading to elimination [12,13]. Combined, evidence from field trials and modelling studies provide strong support for testing the effectiveness of the *Wolbachia* intervention, complemented by other vector control and public health measures, for eliminating dengue as a public health problem in dengue-endemic areas [10].

Establishing that an infectious disease has been eliminated requires different methods than those used for estimating disease frequency (e.g., incidence or prevalence) [14]. For example, while a single confirmed case of a disease is evidence against elimination having been achieved, the absence of detected cases may reflect weaknesses and gaps in the surveillance system, rather than true elimination. For other human infectious diseases for which elimination targets are in place, such as malaria, elimination at national level is certified through a comprehensive independent expert review of disease control and elimination programs, the performance, quality, completeness and output of surveillance systems, a review of disease transmission potential, and assessment of capacity to sustain elimination [15]. These assessments, while rigorous, are frequently qualitative or semi-quantitative in nature, and do not formally quantify the probability that if zero cases have been detected through the public health surveillance system over a period of time, there were truly zero cases in the population in the corresponding period [16].

In contrast to public health, probability-based methods for ascertaining infectious disease absence or elimination are widely used in veterinary epidemiology to substantiate ‘freedom from disease’ status in the context of international trade [17]. One commonly used approach applies scenario tree modelling to estimate the probability that a single case of disease would be detected through surveillance if the disease is present in the population at or above a pre-specified minimum prevalence [16]. This allows estimation of the probability that if zero cases of disease are detected by a surveillance system over time, then the true disease prevalence in the population must be less than the pre-specified minimum – which in practical terms, implies the absence of disease in the population. Unlike survey-based methods, scenario tree modelling allows for estimation of the probability of elimination using data collected through the existing surveillance system (or with planned enhancements to support the demonstration of disease elimination), and estimation of the relative contribution of different surveillance activities to demonstrating elimination. Furthermore, this approach can be used quantitatively to plan an elimination program, including estimation of the number of time periods of zero detected cases required to reach an acceptable level of confidence that elimination has been achieved, as was recently applied to planning for elimination of a livestock disease in New Zealand [18]. Scenario tree-based methods have previously been applied to ascertain the absence or elimination of infectious diseases in human populations, including evaluations of poliovirus surveillance in Australia [19] and the United Kingdom [20]. These methods are also being explored in malaria elimination settings [21].

The aim of this study was to develop a scenario tree modelling framework to guide planning for the potential elimination of locally acquired dengue in Yogyakarta city, Indonesia. The feasibility of elimination of dengue as a public health problem through city-wide deployment of *Wolbachia* intervention, complemented by other vector control measures and enhanced clinical surveillance, will be tested in Yogyakarta city, Indonesia, as part of a future study.

## Methods

### Study design

The study design assumes that after full implementation of the *Wolbachia* intervention and other dengue control measures, there will be a period of time during which zero locally-acquired dengue cases are detected through surveillance, which may be indicative of interruption of local dengue transmission. A scenario tree modelling approach was used to estimate the sensitivity of the dengue surveillance system and the time required to demonstrate elimination of locally-acquired dengue in Yogyakarta city, should the reported incidence of dengue decrease to zero in the future.

### Study setting

Yogyakarta city is the capital city of Yogyakarta province, on the island of Java in Indonesia. It has an estimated population size of 373,589 inhabitants [22]. The healthcare system in Yogyakarta city comprises the public healthcare system, including primary healthcare centres (‘puskesmas’) and public hospitals, and the private healthcare sector. Public and private hospitals are required to notify suspected or confirmed dengue cases to the national surveillance system, via the Yogyakarta District Health Office (DHO). Dengue is suspected based on a clinical case definition for dengue fever (DF) or dengue haemorrhagic fever (DHF; ICD-10 codes A90 [DF] and A91 [DHF]; International Classification of Diseases 10th revision). At present, some suspected cases are tested for dengue virus infection (using immunochromatographic rapid diagnostics tests that detect DENV NS1 antigen and IgM/IgG antibodies as part of routine clinical management or RT-qPCR or NS1 antigen ELISA through participation in research); most are diagnosed and notified based on clinical presentation only.

Between 2006 and 2016, the incidence of dengue case notifications averaged 211 per 100,000 population per year in Yogyakarta [23]. These estimates are likely to considerably underestimate the true incidence of dengue in the population, given the potential for incomplete compliance with reporting requirements, ambulatory cases are not recorded [24], and prior to 2017, only dengue haemorrhagic fever cases, not all dengue cases, were notifiable (i.e. low surveillance system sensitivity). Additionally, an unknown proportion of clinically diagnosed cases are likely to be false positives, as dengue symptoms are similar to other endemic febrile infectious diseases (i.e. low surveillance system specificity).

### Description of dengue elimination intervention

*w*Mel-infected *Ae. aegypti* mosquitoes have been released throughout all residential areas of Yogyakarta city in a phased approach, beginning in August 2016 with a quasi-experimental trial [25] and concluding in January 2021 with the completion of releases throughout the untreated arm of the Applying Wolbachia to Eliminate Dengue (AWED) cluster randomised trial (ClinicalTrials.gov NCT03055585) [10]. A prospective post-implementation study commenced in January 2023 and will run for up to three years, to evaluate the long-term public health impact of citywide *Wolbachia* deployments and the potential elimination of local dengue transmission in Yogyakarta city. A ‘dengue case’ will be defined as an individual with symptomatic, virologically-confirmed dengue infection, resident in Yogyakarta city and without a documented recent travel history outside of Yogyakarta. The case definition does not include asymptomatic dengue virus infections given that i) elimination of dengue ‘as a public health problem’ is the relevant goal rather than complete interruption of transmission; ii) no routine surveillance is in place or planned for asymptomatic infections; and iii) any chains of transmission associated with asymptomatic infections would be expected to lead to at least one symptomatic dengue case, which could then be detected through surveillance.

Recognising the limitations of the sensitivity and specificity of routine hospital-based dengue surveillance, this post-implementation study includes enhanced clinic-based surveillance for dengue among patients presenting with acute febrile illness, to support the demonstration of dengue elimination. Thirteen of the 18 puskesmas in Yogyakarta were selected to participate in the enhanced clinic-based surveillance based on enrolment data from a previous cluster randomised trial [10], and are estimated to account for approximately 85% of all puskesmas presentations for acute febrile illness. All patients aged 3 – 45 years and currently residing in Yogyakarta city who present to these puskesmas with an acute febrile illness of 1– 4 days duration and no localising signs suggestive of a specific diagnosis will be invited to enrol, and a venous blood sample and demographic data will be collected from consenting participants. Patient samples will be tested for the presence of dengue virus using RT-qPCR and NS1 antigen ELISA in a research laboratory, and patients positive by PCR and/or NS1 ELISA will be classified as virologically-confirmed dengue cases. Puskesmas and other outpatient health facilities do not currently routinely report dengue cases to the DHO surveillance system, but in the enhanced clinic-based surveillance study a custom-built data management system will record enrolment data including patients’ residential address, and laboratory diagnostic results. In the immediate term, no change will be made to the existing hospital-based dengue surveillance system. However, as part of a future elimination program, resources would be reallocated to enable follow-up and virological confirmation of each notified dengue case to reduce the false positive rate to zero. The potential for infection to have been acquired outside Yogyakarta city will be assessed in the enhanced clinic-based surveillance by recording at enrolment whether the patient had travelled outside the city in the past 10 days, and in the DHO hospital-based surveillance from a field in the case notification form that records recent travel history. While not a highly accurate measure – and currently recorded incompletely in the DHO surveillance – it is assumed that in an elimination context, detection of suspected locally-acquired dengue would lead to further investigations to resolve the case status and determine whether elimination has likely been achieved.

### Scenario tree modelling framework

A scenario tree is a schematic tree structure of nodes and branches which can be used to describe and assist in the analysis of complex surveillance systems, whilst accounting for heterogeneities in the population risk structure. Risk nodes represent the probability of infection in different subgroups. Detection nodes represent key events, actions, or tests that contribute to an infected individual’s detection in the surveillance process (e.g., that a person attends a clinic; a sample is collected; or that a positive test result is notified). Node probabilities can be estimated from data where available, or informed by expert judgement. The tree structure depicts the various pathways by which an infected individual in the population may be detected, and defines the probabilities associated with each step in the detection process [16,26].

A scenario tree model was developed to describe and analyse the feasibility of demonstrating the elimination of locally acquired dengue in Yogyakarta city, using the existing hospital-based surveillance component supplemented with an enhanced primary care surveillance component (Figure 1). The model includes age and gender as risk factors for being a symptomatic dengue case. Detection nodes include the probability of first seeking care at different types of health facilities (public hospital, puskesmas participating in enhanced dengue surveillance, or other/no healthcare seeking), probability of sample collection, probability of sample testing, diagnostic test sensitivity, and probability of case recording and notification.

**Figure 1:**
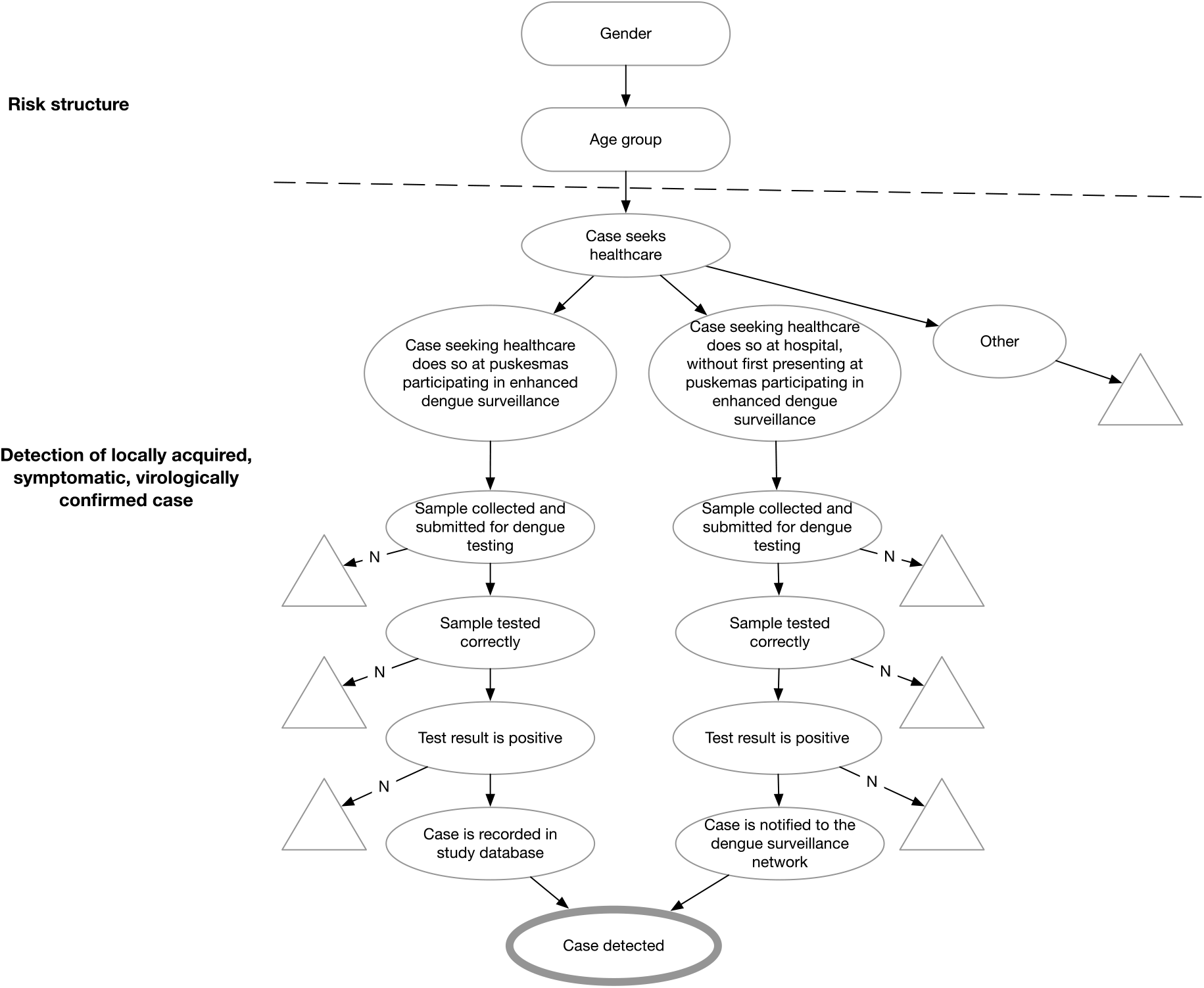
Scenario tree model for the detection of locally acquired, symptomatic, virologically confirmed dengue cases through the public health surveillance system in Yogyakarta city, Indonesia. The scenario tree model represents the planned structure of the surveillance system in the context of a prospective dengue elimination study. This includes an enhanced clinic-based surveillance component at primary health facilities (puskesmas), as well as strengthening the hospital-based surveillance system through introduction of virological testing for all suspected dengue cases. The rectangle nodes represent risk nodes: the probability of being a symptomatic dengue case varies by age and gender. The oval nodes represent detection nodes: key events, actions, or tests that contribute to an infected individual’s detection in the surveillance process. The triangular nodes represent pathways by which cases escape detection.

#### Data sources and model parameterisation

##### Risk nodes

Population data for Yogyakarta city, stratified by gender and age, were obtained from the 2020 census data [20] (Table 1). Relative risks of locally acquired symptomatic dengue were estimated by gender and age group based on data from the AWED trial, supplied by the study authors (Table 1).

**Table 1:**
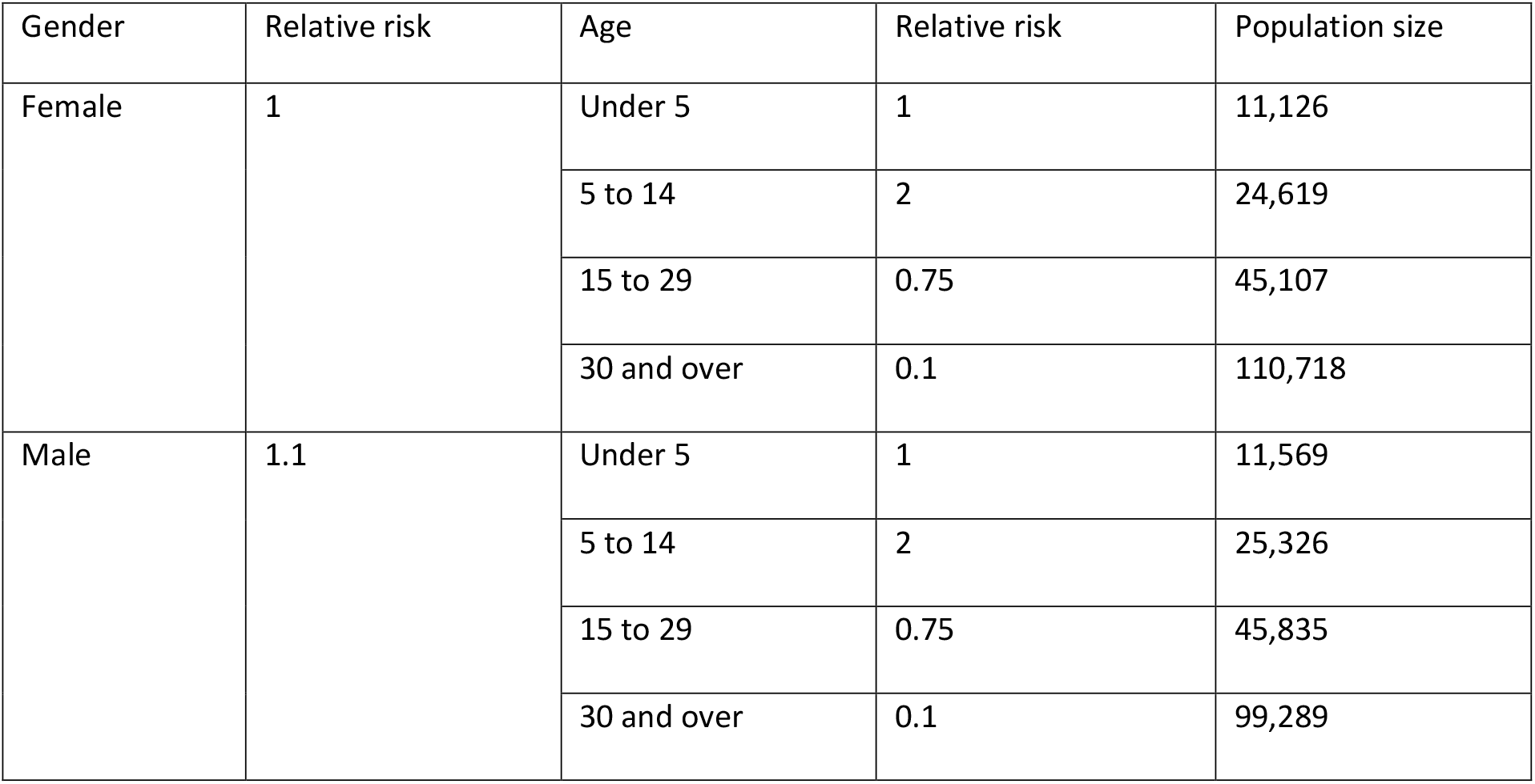
Population level risk factors for locally acquired symptomatic dengue, including estimated relative risk values and population size in each risk stratum.

##### Detection nodes

Parameter distributions were derived from data collected as part of the AWED trial in Yogyakarta city, or estimated by expert opinion if no other data sources were available, as specified. Experts consulted included local clinicians and researchers working on the AWED trial [10,25], who were familiar with the dengue surveillance system in Yogyakarta city.

The probability of seeking healthcare was stratified by age group (Table 2). There was no evidence from AWED trial data to support stratification of healthcare seeking by gender. Other case detection factors (from sampling to case notification) were assumed to be affected by whether a case first seeks healthcare at a primary care clinic participating in the enhanced dengue surveillance program or first seeks healthcare at hospital. Cases seeking healthcare were estimated to be three times more likely to present at clinics participating in enhanced dengue surveillance, compared to seeking healthcare at a hospital (without having attended a clinic participating in the enhanced dengue surveillance program). Approximately 80% of dengue patients seeking healthcare at clinics participating in enhanced dengue surveillance are expected to consent to blood sampling for dengue testing, based on AWED data. In contrast, clinically-diagnosed dengue patients in hospitals were judged to be half as likely to be sampled for dengue testing, given the context of routine clinical management. The coordination of primary care clinic-based enhanced dengue surveillance by dedicated study staff and testing at a research laboratory means that all collected samples are expected to be tested and have results recorded in the study database, whereas there may be some gaps in these steps within the routine hospital surveillance system. Samples collected at clinics participating in enhanced dengue surveillance will be tested according to a using a highly sensitive combination of RT-qPCR and dengue NS1 ELISA, compared to the less sensitive NS1 ELISA or rapid diagnostic test (RDT) used in hospital laboratories, with sensitivity values for NS1 ELISA and RDTs derived from validation studies [27,28]. RT-qPCR is considered the gold-standard reference assay, and was assigned a sensitivity of 0.95 to allow for infrequent DENV infections with a viral load below the limit of detection (Table 2).

**Table 2:**
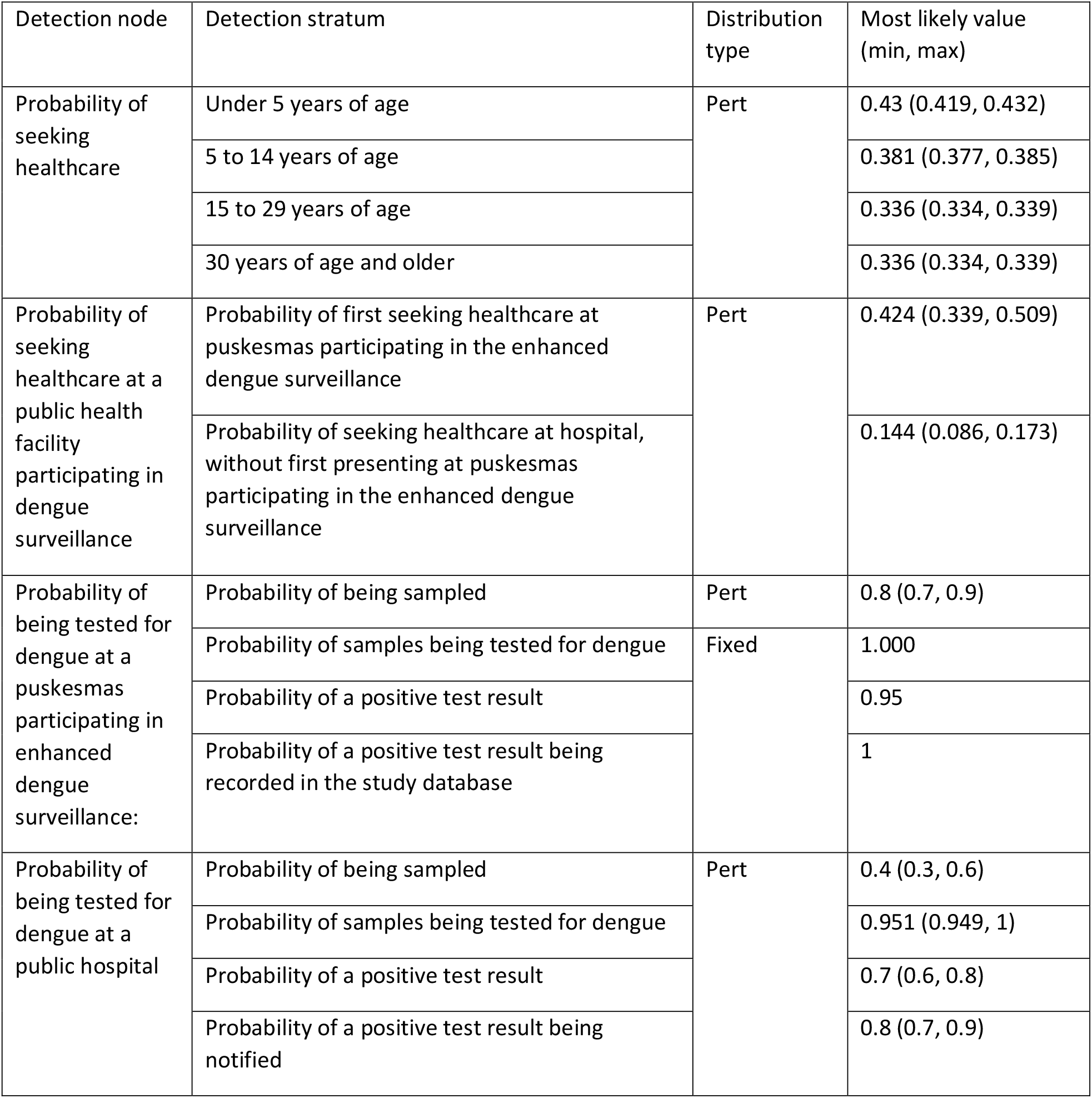
Parameter values for factors affecting the detection of locally acquired clinical dengue infection.

| Detection node | Detection stratum | Distribution type | Most likely value (min, max) |
| --- | --- | --- | --- |
| Probability of seeking healthcare | Under 5 years of age | Pert | 0.43 (0.419, 0.432) |
|  | 5 to 14 years of age |  | 0.381 (0.377, 0.385) |
|  | 15 to 29 years of age |  | 0.336 (0.334, 0.339) |
|  | 30 years of age and older |  | 0.336 (0.334, 0.339) |
| Probability of seeking healthcare at a public health facility participating in dengue surveillance | Probability of first seeking healthcare at puskesmas participating in the enhanced dengue surveillance | Pert | 0.424 (0.339, 0.509) |
|  | Probability of seeking healthcare at hospital, without first presenting at puskesmas participating in the enhanced dengue surveillance |  | 0.144 (0.086, 0.173) |
| Probability of being tested for dengue at a puskesmas participating in enhanced dengue surveillance: | Probability of being sampled | Pert | 0.8 (0.7, 0.9) |
|  | Probability of samples being tested for dengue | Fixed | 1.000 |
|  | Probability of a positive test result |  | 0.95 |
|  | Probability of a positive test result being recorded in the study database |  | 1 |
| Probability of being tested for dengue at a public hospital | Probability of being sampled | Pert | 0.4 (0.3, 0.6) |
|  | Probability of samples being tested for dengue |  | 0.951 (0.949, 1) |
|  | Probability of a positive test result |  | 0.7 (0.6, 0.8) |
|  | Probability of a positive test result being notified |  | 0.8 (0.7, 0.9) |

Parameter inputs for each step in the detection cascade were estimated as either fixed values or as pert distributions (minimum, most likely and maximum values) (Table 2). Pert distributions are considered useful for incorporating expert opinion into a model, where there are inadequate data to parameterise a distribution [24].

#### Surveillance sensitivity

The scenario tree model was firstly used to calculate the sensitivity of the surveillance system. Surveillance system sensitivity is defined as the probability that at least one dengue case would be detected through dengue surveillance if dengue cases are present in the population at a given prevalence, known as the ‘design prevalence’. Design prevalence was set at a single dengue case in the population and converted to a proportion (P^*^) of the total population.

Model calculations used or adapted standard formulae as described by [16] and subsequent methodological developments [18,26,29,30], as follows:

##### Adjusted risk

For each successive risk factor, adjusted risks (*AR*) for each risk group were calculated as:

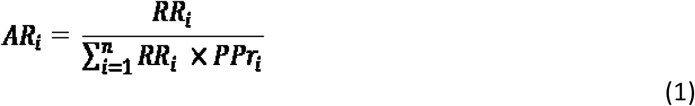

Where *RR*_*i*_ is relative risk for the *i*-th risk category and *PPR*_*i*_ is the population proportion of the *i*-th risk category, for the preceding risk-group in the scenario tree. Thus, for gender, population proportions are for the whole population and for age category they are for the respective genders.

##### Effective probability of infection

Effective probability of infection (*EPI*) was calculated for each risk stratum representing all eight combinations of gender and age category as the product of the respective adjusted risks for each risk factor and the design prevalence (P*):

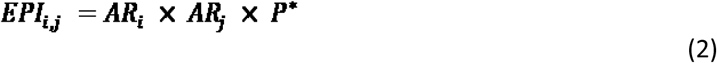

Where *P*^*^ is the design prevalence and *Ar*_*i*_ and *AR*_*j*_ are adjusted risks for the relevant levels of gender and age category, respectively.

##### Unit sensitivity

Unit sensitivity (probability that a dengue case would be detected if they were in the respective risk stratum) was estimated for each of the eight risk strata as the product of the relevant detection probabilities for a person in that stratum.

##### Component and System sensitivity

Component sensitivities for puskesmas and hospitals were calculated separately as:

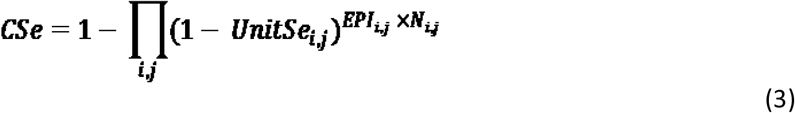

Where *N*_*i,j*_ is the population size for the *i*-th gender and *j*-th age category and *UnitSe* is the unit sensitivity for the relevant stratum for puskesmas and hospital, respectively.

Overall system (population) sensitivity (*SSe*) was then calculated as:

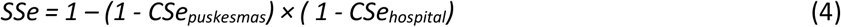

#### Confidence in elimination

Secondly, the scenario tree model allows for testing the hypothesis that locally-acquired dengue has been eliminated, if a period of time passes during which zero cases are detected through surveillance. The null hypothesis was that dengue is present at or above the design prevalence. The alternative hypothesis was that dengue cases, if present, occur at a prevalence lower than the design prevalence. A suitably low design prevalence is chosen such that the inference can be made that if dengue occurs at a prevalence lower than the design prevalence, then dengue has been eliminated as a ‘public health problem’. The level of evidence for the alternative hypothesis is a function of the sensitivity of the surveillance system (*SSe)*, the number of successive time periods with zero reported cases, and the risk of (re-)introduction of dengue into the population in each time period.

Confidence of elimination (*PFree*) was calculated for each time period in two steps. Firstly, a discounted prior confidence (*Prior*_*t*_) was calculated based on the confidence of elimination for the previous time period (*PFree*_*t*-1_), discounted for the probability that locally acquired dengue recurred during the time period (*PRecur*):

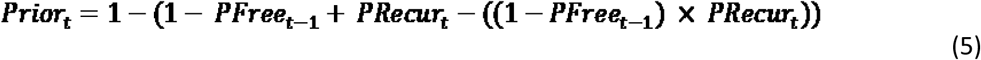

Noting that for the first time period, *PFree*_*t*-1_ was set at 0.5, indicating an uninformed opinion. The probability of recurrence is the probability that one or more cases of locally acquired symptomatic dengue will occur during a time period, in a population in which dengue cases had previously been eliminated. Recurrence of local dengue transmission after prior elimination may occur due to an imported dengue case leading to local transmission. Three separate values (1%, 2% & 5%) for probability of recurrence per month were modelled as a sensitivity analysis to reflect the risk of sporadic imported dengue cases initiating local transmission chain in a vector population with stable *w*Mel introgression, and hence suppressed dengue transmission potential.

Finally, *PFree*_*t*_ was calculated as:

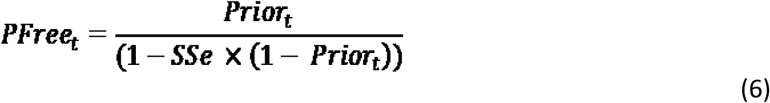

*SSe* was assumed to be the same across all time periods, because it is a probabilistic calculation for future planning purposes and not based on the actual number of possible dengue cases detected by the surveillance system each month. For interpretation, an estimated 80% or higher probability of dengue elimination was considered an acceptable level of evidence for the purposes of ascertaining dengue elimination for this framework.

#### Sensitivity analyses

Several separate simulations were compared to explore the potential impacts of changes in key model parameters, including a reduction in number of primary care clinics participating in enhanced surveillance, as well as modest but realistic improvements to different steps in the detection cascade in clinics and hospitals. The simulations are described in Supplementary Table 1.

#### Model implementation

The model was developed and implemented in the R software environment, version 4.0.3 [31], including the R packages epiR [32], mc2d [33], gplots [34] and RColorBrewer [35]. The time step for the analysis was one month, consistent with the reporting frequency for the dengue surveillance system in Yogyakarta city. The analysis was run for 60 time steps (60 months; 5 years). As described, key inputs were entered as probability distributions and the model was run for 10,000 iterations to generate estimates of system sensitivity per month and confidence of dengue elimination in the population over 60 simulated months.

### Ethics

As the development of the scenario tree modelling framework did not require collection of new data from research participants, ethical approval for the development of the scenario tree modelling framework was not sought. The AWED cluster randomised trial of the *Wolbachia* intervention, which was the source of some aggregate (non-identifiable) data used to parameterise the model, and the clinic-based enhanced surveillance study of dengue elimination in Yogyakarta, in which the framework will be tested, have been reviewed and approved by the institutional review boards of Universitas Gadjah Mada and Monash University.

### Patient and Public Involvement statement

As this study involved the development of a modelling framework based on existing data sources, and did not require the collection of new data from research participants, patients were not involved in the development of the research question, study design or outcome measures. The outcomes of this work will be used to evaluate the evidence for the elimination of dengue in Yogyakarta in 2023-2025, the findings of which will be disseminated to health professionals and the public in Yogyakarta via the Yogyakarta district health office.

## Results

### Unit sensitivity

Median unit sensitivity estimates varied between age group strata, from 0.108 to 0.138 for primary care clinic presentations and 0.013 to 0.016 for hospital presentations per month. Higher values were estimated for the under 5 age group, with the lowest values in the 15 to 29 and over 30 age groups (Supplementary Table 2).

### Surveillance system sensitivity

For the default simulation, median surveillance system sensitivity was 0.131 (95% PI 0.111 – 0.152) per month. Median component sensitivities were 0.118 (95% PI 0.099 – 0.139) per month for clinics participating in the enhanced dengue surveillance program and 0.014 (95% PI 0.010 – 0.020) per month for public hospital surveillance (Figure 2).

**Figure 2:**
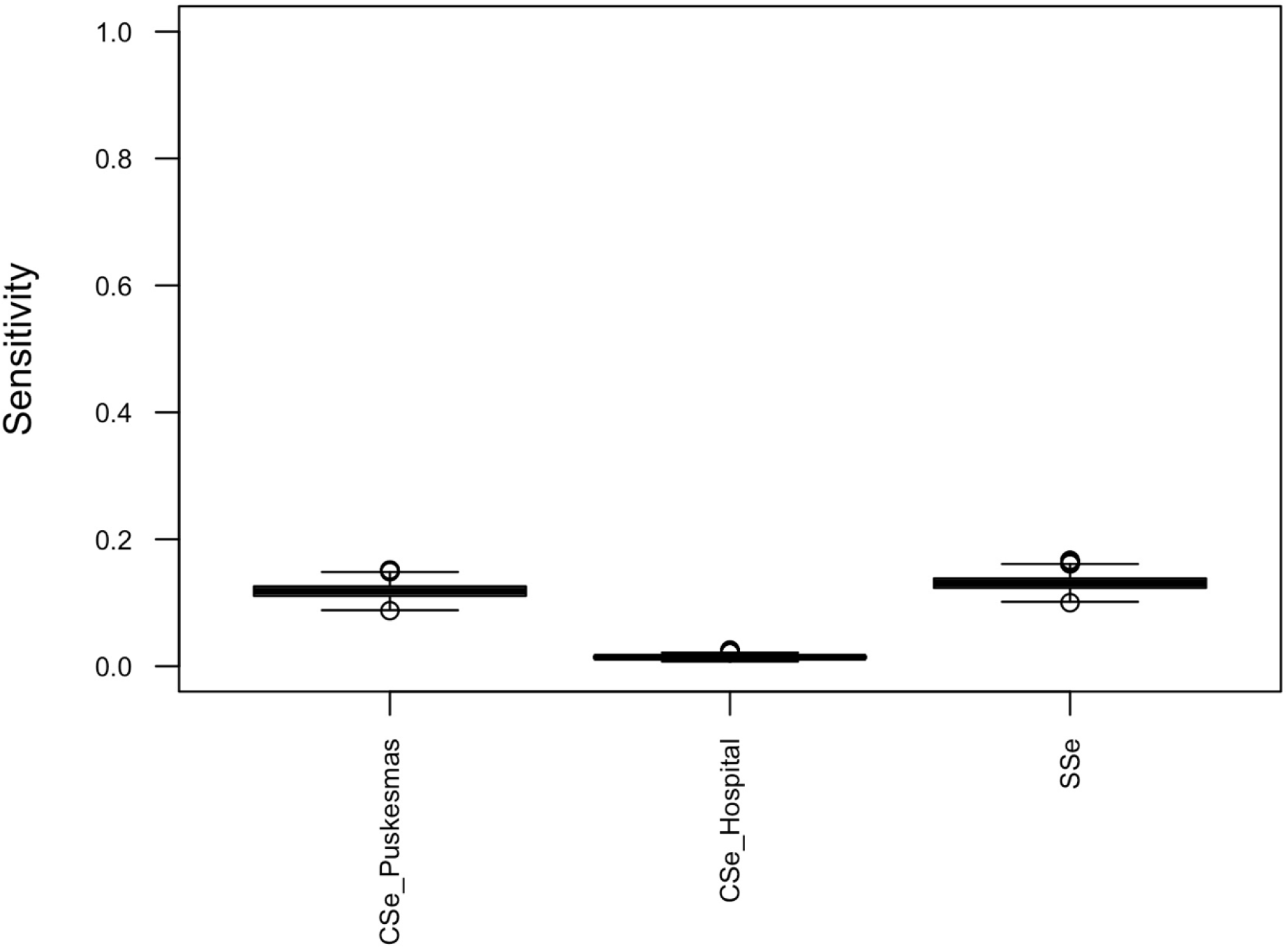
Box and whisker plot of component sensitivities for puskesmas-based enhanced dengue surveillance (CSe_puskesmas) and hospital surveillance for dengue (CSe_hospital), and overall surveillance system sensitivity for dengue cases (SSe), per month.

The alternative simulations investigated produced relatively small changes in median system sensitivity (Supplementary Table 1). The largest change in system sensitivity induced by a change in a single parameter was observed for Simulation 2 (a reduction in probability of presenting to clinics participating in the enhanced surveillance program by 20%), which reduced surveillance system sensitivity from 0.131 to 0.105. Simulation 7 (representing multiple improvements in the surveillance system) resulted in an increase in median system sensitivity to 0.162.

### Confidence of elimination

For a probability of recurrence of local transmission of 1% per month, median confidence in dengue elimination reached 80% after 13 months of zero detected dengue cases and 90% after 25 months, and plateaued at approximately 93% after 44 months (Figure 3). For a probability of recurrence of 2%, median confidence of dengue elimination of 80% was reached after 19 months, and plateaued at approximately 86% after 42 months. For a probability of recurrence of 5%, median confidence of dengue elimination never reached 80%, plateauing at approximately 64% after 33 months.

**Figure 3:**
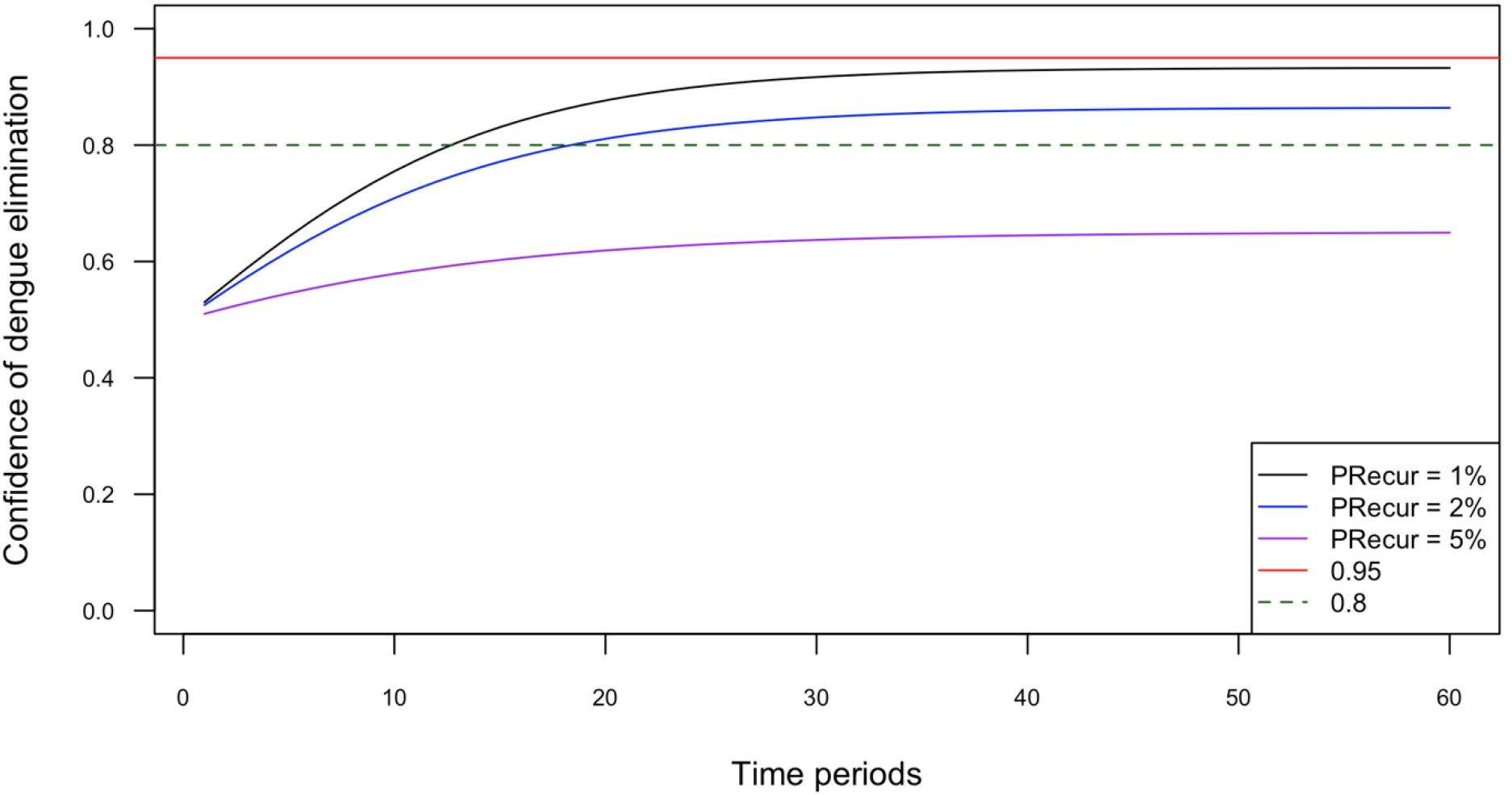
Median confidence of dengue elimination over 60 months, for probability of recurrence (PRecur) of 1%, 2% and 5% per month. PRecur: probability of recurrence of local dengue transmission in a single time period (one month). Reference lines for level of confidence of elimination are plotted at 80% confidence (green dashed line), and 95% confidence (red reference line).

The sensitivity analyses mostly resulted in minor changes in the trajectory of confidence of elimination over time (Supplementary Figure 1). However, for Simulation 2 (reduced number of clinics participating in enhanced surveillance), the median confidence in dengue elimination reaching 80% was extended to approximately 18 months of zero reported cases; whereas in Simulation 7 (a combination of modest improvements to the surveillance system), it was reduced to within 10 months.

## Discussion

### Feasibility of demonstrating dengue elimination in Yogyakarta city

This study suggests that with a combination of hospital-based surveillance and enhanced primary care clinic-based surveillance for dengue, an acceptable level of confidence (80% probability) in the elimination of locally acquired dengue can be reached within 2 years. Potentially, it could be reached in as few as 13 months, as long as the high *Wolbachia* coverage and virus-blocking activity is sustained such that the risk of resumption of local transmission after a period of local elimination is 1% or lower. This scenario is realistic because elimination of local dengue transmission would not be plausible if the true probability of recurrence of local transmission arising from an imported case in an otherwise dengue-free population was as high as 5% per month—this probability would be inconsistent with stable suppression of local transmission through the *Wolbachia* intervention. Of note, these results suggest that pragmatic demonstration of the elimination of locally-acquired dengue can be reached in a shorter timeframe than the alternative approach of convening an independent expert review panel, as commonly used for other infectious disease elimination programs [15]. However, it must be noted that given the surveillance components and detection node parameter values included in the model, it would not be possible to reach 95% confidence in elimination of locally-acquired dengue. This more stringent threshold has been proposed where demonstration of infectious disease elimination is used to guide decision-making about cessation of disease control activities [14]. In Yogyakarta city, it is unlikely that dengue control activities would be halted even if elimination appears to have been reached, as the city remains susceptible to imported cases.

Increasing the surveillance system sensitivity could shorten the time to first ascertainment of elimination of dengue and increase the level of confidence in elimination. The nodes in the scenario tree with the lowest probability estimates represent the greatest opportunities to have a substantial impact through interventions. In addition to modest improvements to surveillance data collection and reporting modelled as part of the sensitivity analysis (Supplementary Table 1), surveillance system sensitivity could be further improved through increasing the probability of care-seeking at clinics participating in enhanced surveillance, which would require a comprehensive community engagement strategy as part of a dengue elimination program.

A key assumption of scenario tree modelling is that surveillance system specificity is 100%. This does not imply that diagnostic tests for dengue must be 100% specific – rather, that in an elimination context, all test-positive cases will be further investigated to resolve their status as true positive or false positive cases. For dengue, this would include follow-up virological testing for all clinically diagnosed cases, as well as reliably distinguishing local from imported cases through travel history questionnaires and other sources of epidemiological and entomological data. Though challenging to consistently implement in practice, especially in lower resource settings, this approach reflects how suspected cases for other infectious diseases that are believed to have been eliminated are investigated, such as poliovirus [20]. Therefore, the scenario-tree modelling framework presented here is relevant for a dengue elimination program with adequate resources allocated to follow-up and resolve the status of all suspected dengue cases.

### Surveillance system sensitivity for other human infectious diseases

The dengue surveillance system sensitivity in Yogyakarta city, with the planned addition of enhanced clinic-based surveillance, was estimated at 0.131 (95% PI 0.111, 0.152) per month. As the use of scenario tree modelling to estimate confidence in elimination or absence of disease in human populations is an emerging methodology, it is difficult to directly compare the findings of this study to others in the literature. For example, the estimated median monthly sensitivity of clinical surveillance for wild poliovirus infection amongst children aged less than fifteen years in Australia was estimated at 0.082 (95% PI 0.053, 0.121) [19]. In the United Kingdom, clinical poliovirus surveillance system sensitivity was estimated at 0.00324 (95% CI 0.00177 – 0.00481) per month for wild-type poliovirus and 0.000336 (95% CI 0.000160 – 0.000641) per month for vaccine-derived poliovirus infection [20]. However, in these two studies, the case definition referred to any poliovirus infection, including asymptomatic infection, whereas the dengue framework adopts a more pragmatic definition of elimination focused on clinical cases.

Another study applied the scenario tree modelling approach to estimate the sensitivity of surveillance for leptospirosis in Ecuador, where leptospirosis is endemic [36]. This study estimated surveillance sensitivity at substantially higher design prevalence than used here (ranging from 1% to 30% across three geographic regions), consistent with the goal of monitoring an endemic disease rather than demonstrating elimination. This study reported surveillance system sensitivities ranging from 0.29 (95% PI 0.02 – 0.82) to 0.85 (95% PI 0.41 – 0.99). As surveillance sensitivity is defined as the probability of detecting at least one case, higher surveillance system sensitivities are expected as disease prevalence increases.

A recent study aimed to establish a framework for estimating surveillance sensitivity for clinical malaria cases in the context of a malaria elimination target in two districts in Indonesia, including one district in Yogyakarta province [37]. This study estimated parameters for the malaria case detection cascade from survey and interview data in the study area, but did not present an overall estimate of surveillance system sensitivity. Estimates of the probability of seeking care averaged 0.09 (± 0.05 sd) and the probability of testing averaged 0.16 (± 0.29 sd). The lower care-seeking estimates compared to the present study likely reflect disparities in access to health care between urban residents in Yogyakarta city and rural residents in malaria-endemic areas, with care seeking in the malaria study related to availability of antimalarial drugs, antimalarial drug stockouts, and travel time to health facilities [37].

### Strengths and limitations

This is the first study to establish a framework for planning and demonstrating dengue elimination, with parameter inputs that can be readily obtained from routinely collected data or expert opinion from program managers. The framework can be adapted to support planning for dengue elimination as evidence-based interventions including new dengue vaccines and *Wolbachia* are implemented at increasing scale. Where current surveillance activities and coverage are inadequate to support demonstrating elimination, the framework highlights key points in the surveillance cascade to target to improve surveillance system sensitivity overall, which may vary across settings.

A limitation of this study is that most parameter inputs (with the exception of age variations in healthcare seeking behaviour) were derived from expert opinion, as no alternative data sources were available. Though considered a suitable method for eliciting parameter values in scenario tree modelling [16], the parameter values derived from expert opinion should be empirically validated as part of the next phase of the dengue elimination research in Yogyakarta city. The model presented herein can then be re-run with empirical data as it becomes available to refine the output estimates.

A second limitation is that the model only considers age and gender as risk factors for dengue. Other sociodemographic risk factors, such as house construction or outdoor occupation, may be relevant to include if population-level data sources become available. Fine-scale spatial risk and detection nodes were considered for inclusion in the model, but the lack of reliable data to distinguish dengue-related risk factors between city districts, and challenges taking cross-city movements into account, resulted in the exclusion of spatial area from the model. This limits the generalisability of the current framework to larger geographic areas, where spatial differences are more pronounced (e.g., differences in healthcare seeking between urban and rural areas).

## Conclusion

The scenario tree modelling framework offers a replicable, robust method for planning and assessing progress towards elimination of dengue as a public health problem following full implementation of the *Wolbachia* intervention and enhanced clinic-based dengue surveillance in Yogyakarta city, Indonesia. The framework can be readily expanded and adapted to other geographic settings where the *Wolbachia* intervention – and in future, dengue vaccines – are implemented to control and eliminate dengue.

## Data Availability

All data relevant to the study are included in the article or uploaded as supplementary information.

## Supplementary information

**Supplementary Table 1:** Summary description of multiple separate simulations conducted

| Simulation | Change from default simulation | New distribution parameters | Median system sensitivity (95% PI)* |
| --- | --- | --- | --- |
| 1 | Default parameters as described | No change | 0.131 (0.111 – 0.152) |
| 2 | Probability of seeking healthcare at puskesmas participating in the enhanced dengue surveillance program reduced proportionally by 20%, reflecting a decrease in the number of puskesmas participating in enhanced surveillance due to resource constraints | pert(0.271, 0.339, 0.407) | 0.105 (0.089 – 0.122) |
| 3 | Probability of first seeking healthcare at hospital (i.e., without first presenting at puskesmas participating in the enhanced dengue surveillance) increased by 5% (absolute) | pert(0.136, 0.194, 0.223) | 0.132 (0.112 – 0.153) |
| 4 | Probability of sampling at puskesmas participating in the enhanced dengue surveillance program increased, reflecting increased awareness of the dengue elimination goal amongst healthcare providers | pert(0.8, 0.9, 0.95) | 0.142 (0.121 – 0.163) |
| 5 | Probability of sampling at hospital increased, reflecting increased awareness of the dengue elimination goal amongst healthcare providers | pert(0.7, 0.8, 0.9) | 0.138 (0.118 – 0.159) |
| 6 | Probability of notification of a positive result at a hospital increased, reflecting increased awareness of the dengue elimination goal amongst healthcare providers | pert(0.9, 0.95, 0.98) | 0.130 (0.110 – 0.151) |
| 7 | Changes per simulations 3 – 6 combined in a single model |  | 0.162 (0.142 – 0.184) |

Table notes: \*Assumes a probability of recurrence of local dengue transmission of 1% per month.

**Supplementary Table 2:** Unit sensitivities (USe) by age, gender, and type of health facility

|  | Median USE (5th-95th percentile) |  |
| --- | --- | --- |
| Risk stratum | Puskesmas | Hospital |
| Under 5_female | 0.138 (0.118-0.159) | 0.016 (0.012-0.022) |
| 5 to 14_female | 0.123 (0.105-0.141) | 0.015 (0.011-0.02) |
| 15 to 29_female | 0.108 (0.093-0.125) | 0.013 (0.009-0.017) |
| 30 and over_female | 0.108 (0.093-0.125) | 0.013 (0.009-0.017) |
| Under 5_male | 0.138 (0.118-0.159) | 0.016 (0.012-0.022) |
| 5 to 14_male | 0.123 (0.105-0.141) | 0.015 (0.011-0.02) |
| 15 to 29_male | 0.108 (0.093-0.125) | 0.013 (0.009-0.017) |
| 30 and over_male | 0.108 (0.093-0.125) | 0.013 (0.009-0.017) |

**Supplementary Figure 1:**
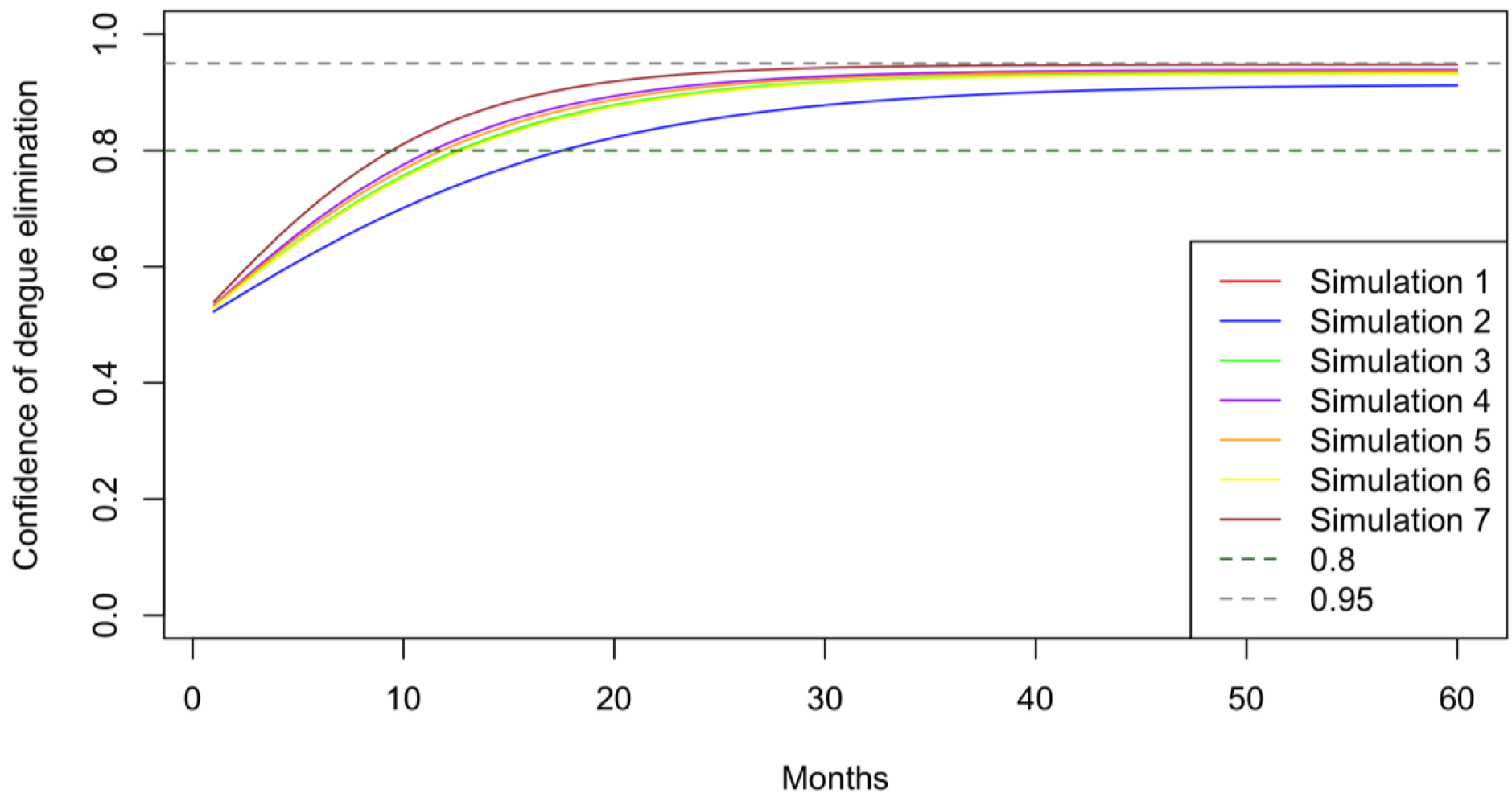
Median confidence of elimination over 60 months for seven alternative simulations and assuming a probability of recurrence of 1% per month.

## References

1 Simmons CP, Farrar JJ, van Vinh Chau N, et al. Dengue. N Engl J Med 2012;366:1423–32. doi:10.1056/NEJMra1110265

2 World Health Organization. Dengue and severe dengue. 2022.https://www.who.int/news-room/fact-sheets/detail/dengue-and-severe-dengue (accessed 13 Jun 2022).

3 Stanaway JD, Shepard DS, Undurraga EA, et al. The global burden of dengue: an analysis from the Global Burden of Disease Study 2013. Lancet Infect Dis 2016;16:712–23. doi:10.1016/S1473-3099(16)00026-8

4 World Health Organization. Ten health issues WHO will tackle this year. 2019.https://www.who.int/news-room/spotlight/ten-threats-to-global-health-in-2019 (accessed 13 Jun 2022).

5 Bian G, Xu Y, Lu P, et al. The endosymbiotic bacterium Wolbachia induces resistance to dengue virus in Aedes aegypti. PLoS Pathog 2010;6:e1000833.

6 Frentiu FD, Zakir T, Walker T, et al. Limited dengue virus replication in field-collected Aedes aegypti mosquitoes infected with Wolbachia. PLoS Negl Trop Dis 2014;8:e2688.

7 Hoffmann AA, Montgomery BL, Popovici J, et al. Successful establishment of Wolbachia in Aedes populations to suppress dengue transmission. Nature 2011;476:454–7.

8 Lambrechts L, Ferguson NM, Harris E, et al. Assessing the epidemiological effect of Wolbachia for dengue control. Lancet Infect Dis 2015;15:862–6.

9 Walker T, Johnson PH, Moreira LA, et al. The wMel Wolbachia strain blocks dengue and invades caged Aedes aegypti populations. Nature 2011;476:450–3.

10 Utarini A, Indriani C, Ahmad RA, et al. Efficacy of Wolbachia-infected mosquito deployments for the control of dengue. N Engl J Med 2021;384:2177–86.

11 Ryan PA, Turley AP, Wilson G, et al. Establishment of wMel Wolbachia in Aedes aegypti mosquitoes and reduction of local dengue transmission in Cairns and surrounding locations in northern Queensland, Australia. Gates Open Res 2019;3:1547. doi:10.12688/gatesopenres.13061.2

12 Ferguson NM, Hue Kien DT, Clapham H, et al. Modeling the impact on virus transmission of Wolbachia-mediated blocking of dengue virus infection of Aedes aegypti. Sci Transl Med 2015;7:279ra37–279ra37. doi:10.1126/scitranslmed.3010370

13 O’Reilly KM, Hendrickx E, Kharisma DD, et al. Estimating the burden of dengue and the impact of release of wMel Wolbachia-infected mosquitoes in Indonesia: a modelling study. BMC Med 2019;17:172. doi:10.1186/s12916-019-1396-4

14 Michael E, Smith ME, Katabarwa MN, et al. Substantiating freedom from parasitic infection by combining transmission model predictions with disease surveys. Nat Commun 2018;9:1–13. doi:10.1038/s41467-018-06657-5

15 Lindblade KA, Li Xiao H, Tiffany A, et al. Supporting countries to achieve their malaria elimination goals: the WHO E-2020 initiative. Malar J 2021;20:1–11. doi:10.1186/s12936-021-03998-3

16 Martin PAJ, Cameron AR, Greiner M. Demonstrating freedom from disease using multiple complex data sources: 1: A new methodology based on scenario trees. Prev Vet Med 2007;79:71–97.

17 Cameron AR, Baldock FC. A new probability formula for surveys to substantiate freedom from disease. Prev Vet Med 1998;34:1–17.

18 Cowled BD, Sergeant ES, Leslie EE, et al. Use of scenario tree modelling to plan freedom from infection surveillance: Mycoplasma bovis in New Zealand. Prev Vet Med 2022;198:105523.

19 Watkins RE, Martin PAJ, Kelly H, et al. An evaluation of the sensitivity of acute flaccid paralysis surveillance for poliovirus infection in Australia. BMC Infect Dis 2009;9:1–13.

20 O’Reilly KM, Grassly NC, Allen DJ, et al. Surveillance optimisation to detect poliovirus in the pre-eradication era: a modelling study of England and Wales. Epidemiol Infect 2020;148.

21 Stresman G, Cameron A, Drakeley C. Freedom from infection: confirming interruption of malaria transmission. Trends Parasitol 2017;33:345–52.

22 Badan Pusat Statistik. Hasil sensus penduduk 2020. Indonesia: 2021. https://www.bps.go.id/pressrelease/2021/01/21/1854/hasil-sensus-penduduk-2020.html

23 Indriani C, Ahmad RA, Wiratama BS, et al. Baseline Characterization of Dengue Epidemiology in Yogyakarta City, Indonesia, before a Randomized Controlled Trial of Wolbachia for Arboviral Disease Control. Am J Trop Med Hyg 2018;99:1299–307. doi:10.4269/ajtmh.18-0315

24 Wahyono TYM, Nealon J, Beucher S, et al. Indonesian dengue burden estimates: review of evidence by an expert panel. Epidemiol Infect 2017;145:2324–9.

25 Indriani C, Tantowijoyo W, Rances E, et al. Reduced dengue incidence following deployments of Wolbachia-infected Aedes aegypti in Yogyakarta, Indonesia: a quasi-experimental trial using controlled interrupted time series analysis. Gates Open Res 2020;4:50. doi:10.12688/gatesopenres.13122.1

26 Martin PAJ, Cameron AR, Barfod K, et al. Demonstrating freedom from disease using multiple complex data sources: 2: case study—classical swine fever in Denmark. Prev Vet Med 2007;79:98–115.

27 Hunsperger EA, Sharp TM, Lalita P, et al. Use of a Rapid Test for Diagnosis of Dengue during Suspected Dengue Outbreaks in Resource-Limited Regions. J Clin Microbiol 2016;54:2090–5. doi:10.1128/JCM.00521-16

28 Hunsperger EA, Yoksan S, Buchy P, et al. Evaluation of Commercially Available Diagnostic Tests for the Detection of Dengue Virus NS1 Antigen and Anti-Dengue Virus IgM Antibody. PLoS Negl Trop Dis 2014;8:e3171. doi:10.1371/journal.pntd.0003171

29 Lyngstad TM, Hellberg H, Viljugrein H, et al. Routine clinical inspections in Norwegian marine salmonid sites: A key role in surveillance for freedom from pathogenic viral haemorrhagic septicaemia (VHS). Prev Vet Med 2016;124:85–95. doi:https://doi.org/10.1016/j.prevetmed.2015.12.008

30 Sergeant ES, Dries LR, Moore KM, et al. Estimating population sensitivity and confidence of freedom from highly pathogenic avian influenza in the Victorian poultry industry using passive surveillance. Prev Vet Med 2022;202:105622.

31 R Core Team. R: A language and environment for statistical computing. 2022.http://www.R-project.org/ (accessed 4 Jun 2022).

32 Stevenson M, Sergeant E. epiR. 2022.http://www.R-project.org/ (accessed 4 Jun 2022).

33 Pouillot R, Delignette-Muller M, Denis J. Tools for Two-Dimensional Monte-Carlo Simulations. 2017.https://cran.r-project.org/package=mc2d (accessed 4 Jun 2022).

34 Warnes G, Bolker B, Bonebakker L, et al. gplots: Various R Programming Tools for Plotting Data version 3.1.3 from CRAN. 2020.https://rdrr.io/cran/gplots/ (accessed 11 Nov 2020).

35 Neuwirth E. ColorBrewer Palettes. 2014.https://cran.r-project.org/package=RColorBrewer (accessed 4 Jun 2022).

36 Calero ML, Monti G. Assessment of the Current Surveillance System for Human Leptospirosis in Ecuador by Decision Analytic Modeling. Front Public Health 2022;10:711938. doi:10.3389/fpubh.2022.711938

37 Ahmad RA, Nelli L, Surendra H, et al. A framework for evaluating health system surveillance sensitivity to support public health decision-making for malaria elimination: a case study from Indonesia. BMC Infect Dis 2022;22:619. doi:10.1186/s12879-022-07581-2

